# Development of a Human Evaluation Framework and Correlation with Automated Metrics for Natural Language Generation of Medical Diagnoses

**DOI:** 10.1101/2024.03.20.24304620

**Authors:** Emma Croxford, Yanjun Gao, Brian Patterson, Daniel To, Samuel Tesch, Dmitriy Dligach, Anoop Mayampurath, Matthew M Churpek, Majid Afshar

**Affiliations:** Department of Medicine, School of Medicine and Public Health, University of Wisconsin Madison; Biostatistics and Medical Informatics, School of Medicine and Public Health, University of Wisconsin Madison; Department of Emergency Medicine, School of Medicine and Public Health, University of Wisconsin Madison; Department of Computer Science, Loyola University Chicago

## Abstract

In the evolving landscape of clinical Natural Language Generation (NLG), assessing abstractive text quality remains challenging, as existing methods often overlook generative task complexities. This work aimed to examine the current state of automated evaluation metrics in NLG in healthcare. To have a robust and well-validated baseline with which to examine the alignment of these metrics, we created a comprehensive human evaluation framework. Employing ChatGPT-3.5-turbo generative output, we correlated human judgments with each metric. None of the metrics demonstrated high alignment; however, the SapBERT score—a Unified Medical Language System (UMLS)-showed the best results. This underscores the importance of incorporating domain-specific knowledge into evaluation efforts. Our work reveals the deficiency in quality evaluations for generated text and introduces our comprehensive human evaluation framework as a baseline. Future efforts should prioritize integrating medical knowledge databases to enhance the alignment of automated metrics, particularly focusing on refining the SapBERT score for improved assessments.

## Introduction

In the rapidly developing field of Natural Language Generation (NLG) evaluation, the advent of Large Language Models (LLMs) has opened unprecedented opportunities for assessing the quality of the generated text, highlighting the capacity for more detailed and nuanced evaluations. Despite this progress, our foundation still heavily relies on traditional metrics such as the Recall-Oriented Understudy for Gisting Evaluation (ROUGE) score. While ROUGE remains a staple for summarization tasks, its dependence on string matching reveals significant limitations, especially in capturing the subtleties of texts with high levels of abstraction, like those encountered in medical diagnostics. Consequently, there is a noticeable disconnect between the capabilities of these current evaluation methodologies and the demands of complex generative tasks, underscoring a persistent gap in our understanding of their alignment.

Within the medical sphere, the assessment of NLG takes on heightened significance because of the need to prevent biases, mitigate potential harm, and ensure accurate diagnoses. Consequently, a pressing need arises for comprehensive and well-validated evaluation methodologies tailored to the unique challenges posed by clinical diagnostic decision support. To navigate these challenges, our primary objective was to examine automated metrics for their alignment with human judgments in clinical domain tasks. This work began with an exploration of the current state of clinical NLG evaluation, emphasizing the necessity for a nuanced approach in the diagnostic generation context. Subsequently, we delved into the critical examination of important metrics, considering their applicability and limitations in the realm of diagnosis generation. Additionally, we introduced a comprehensive human evaluation framework with robust interrater reliability and content validity, designed to assess the quality of each automated evaluation metric. By focusing on the interplay between NLG advancements and diagnostic generation evaluation, this work aimed to contribute valuable insights to the ongoing discourse in the evaluation of generative artificial intelligence for medical purposes.

## Background and Related Work for Automated Metrics

Across the spectrum of NLP tasks, we focused on automated metrics currently available for summarization and question-answering as the tasks most closely aligned with diagnosis generation. A comprehensive literature search was conducted across the Association for Computational Linguistics (ACL) anthology, Medline, and Scopus databases between April 20, 2023, and August 3, 2023, for literature that utilized human or automated evaluation metrics on such tasks. In the screening phase, the following criteria were met: (1) utilization of a human or automated evaluation metric; (2) a task related to text generation from a LLM; (3) a summarization task; and (4) in the clinical domain. The keywords used in the search are listed in Supplementary Materials. Additional literature was pulled that detailed fundamental knowledge on any of the search criteria based upon repeated references or citations from those already being reviewed. A review of the paper abstracts was performed to confirm the inclusion criteria of papers and a full paper review was completed in the final selection. Results were imported into the Zotero citation management software (v6.0.27). The search resulted in 82 papers covering 105 metrics that were included for full review by author EC. The full search workflow is shown in Figure 1.

**Figure 1:**
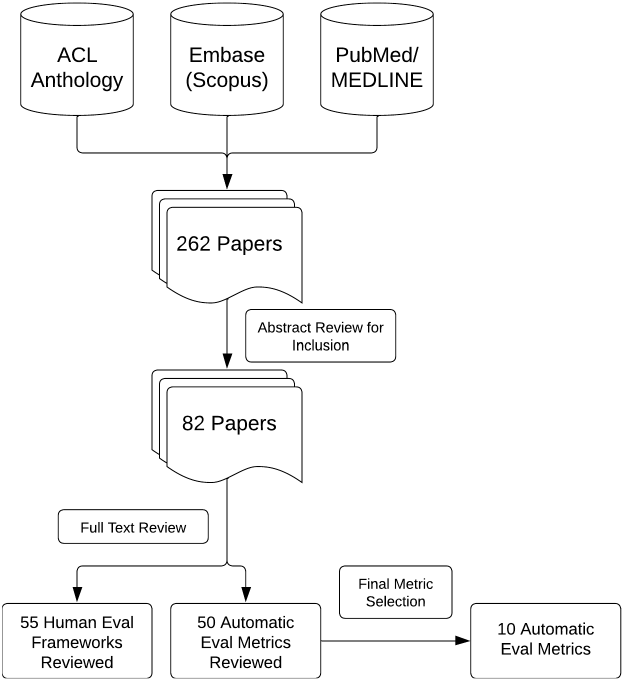
Overview of Literature Review Process and Metric Selection

**Figure 2:**
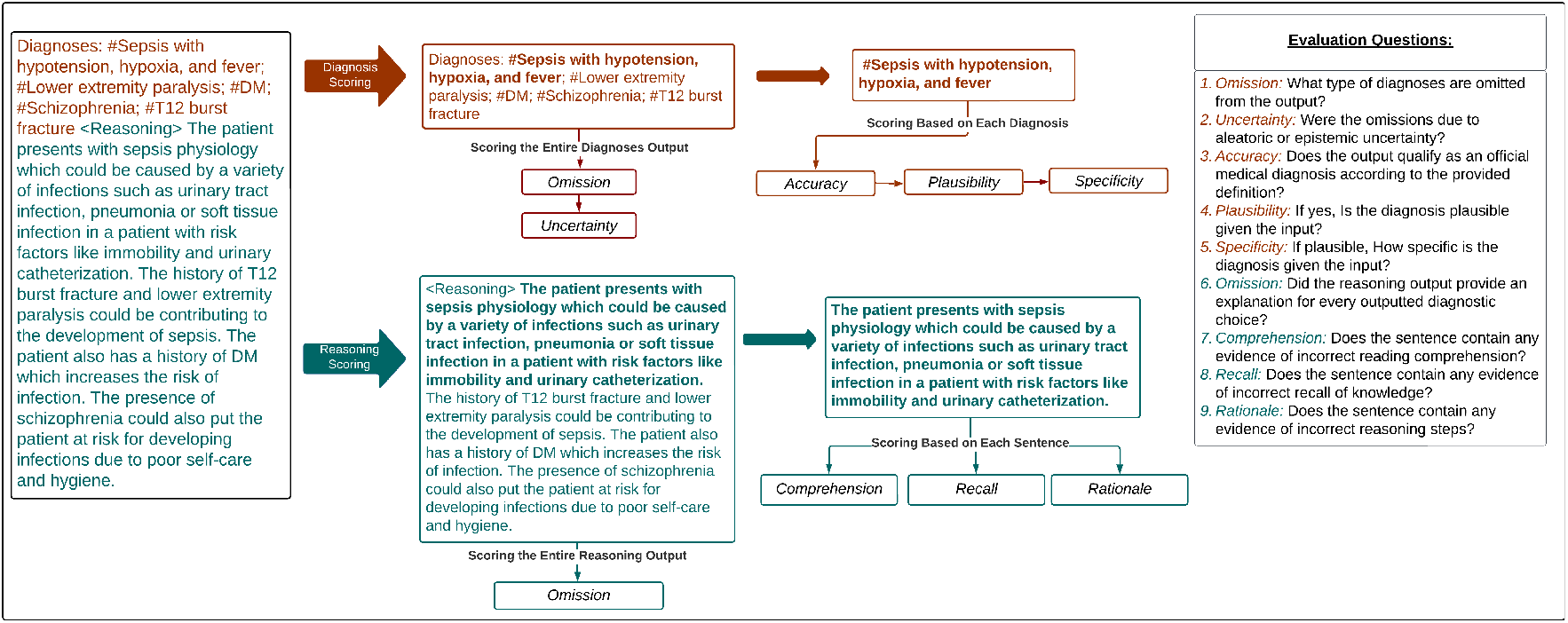
Human Evaluation Survey Workflow. An overview of the human evaluation survey workflow and the questions associated with each component of the evaluation. This framework consists of a Diagnostic Accuracy flow (orange) and a Diagnostic Reasoning flow (teal).

A final set of automated metrics was chosen for inclusion because of their common use in tasks similar to the BioNLP 2023 ProbSum task [1] and/or positive reported results for correlation with human evaluations in the clinical domain. Ten metrics were included: (1) ROUGE-L [2], (2) UMLS Scorer [3], (3) CUI F-Score [4], (4) SapBERT Score [5], (5) clinicalBERT Score [6], (6) PubMedBERT Score [7], (7) clinicalBART Score [8], (8) Commonsense Transformers for automated Knowledge Graph Construction (COMET) [9], (9) ProbSum Trained COMET [9], and (10) clinicalBLEURT [10]. These 10 metrics were classified into four categories based on the nature of the metric: (1) N-gram Overlap Based; (2) UMLS Based; (3) Non-UMLS Embedding Based; and (4) Learned Regression Based.

N-gram overlap metrics were those that compared candidate and reference output using the number of words, phrases, or some other sub-sequence type that were present in the outputs. Our only n-gram overlap metric was the ROUGE-L [2] because it was the current gold standard for evaluating text summarization. ROUGE-L is the longest common sub-sequence variant of ROUGE and uses that common sub-sequence as the n-gram overlap.

The UMLS based metrics were UMLS Scorer, CUI F-Score, and SapBert Score. Each of these relied on the Unified Medical Language System (UMLS) [11] to generate embedding representations of the candidate and reference outputs for comparison. The UMLS Scorer [3] used UMLS-based knowledge graph embeddings [10] and computed an F-score. The CUI F-score [4] also computed an F-score but used Concept Unique Identifiers (CUIs) from the UMLS. Finally, SapBert Score created embeddings using a concept-based model that was trained on the UMLS. This metric is a variation on the BERTScore [12], a metric that computed the maximum pairwise cosine similarity between words in reference and candidate output

The Non-UMLS embedding-based metrics were the clinicalBERT Score, PubMedBERT Score, and clinicalBART Score. The clinicalBERT and PubMedBERT scores are additional variations on the BERTScore where the concept-based model was trained on clinical notes from Medical Information Mart for Intensive Care (MIMIC)-III [13] and abstracts from PubMed, respectively. The clinicalBART Score was a variation on the BARTScore [8] using a Bidirectional and Auto-Regressive Transformer (BART) model fine-tuned on abstracts from PubMed to create embeddings. The score was computed as the sum of the log probabilities of the candidate output given the reference output.

Finally, the Learned Regression-based metrics were COMET, a customized COMET trained on our training data (ProbSum COMET), and clinicalBLEURT. COMET is a neural framework trained for evaluation [9]. We looked at both the default COMET model and fine-tuned an additional COMET model using a small portion of data from the train set of the ProbSum task. Both of these models use a regression approach and were built upon the XLM-RoBERTA pre-trained text encoder. We also included the clinical BLEURT score [10]. This metric is a variation on a learned evaluation BLEU score, called the Bilingual Evaluation Understudy with Representations from Transformers (BLEURT). BLEURT was a regression-based metric that was fine-tuned on clinical notes [14].

## Methods

### Problem Summarization Shared Task for Diagnosis Generation

We utilized data from the Problem List BioNLP Summarization (ProbSum) 2023 Shared Task [1], which was one of the first natural language generation tasks in the medical domain. The goal of this task was to summarize a patient’s active problems/diagnoses given the Subjective, Objective, and Assessment sections from the daily progress notes. The Plan section was used to label the gold standard diagnoses. The progress notes came from MIMIC-III, and the full annotation protocol was detailed previously [4].

### Large Language Model for Diagnosis Generation

We selected ChatGPT-3.5-turbo as our language model because of its superior performance and availability under an exemption status without sharing data [15] [16] [17]. In compliance with the Physionet Credentialed Data Use Agreement, Azure OpenAI service approved our request to opt out of human review in our institution’s Azure environment to run ChatGPT. For prompt design in the task, ChatGPT was asked to paraphrase 50 prompts from a prompt initially authored by human subject matter experts. The lowest perplexity prompt was chosen [18] and self-consistency [19] metrics were also performed to achieve the final, optimal prompt.

The final prompt in the persona of a physician was the following: “Imagine you are a medical professional, and generate the top three direct and indirect, differential diagnoses from the input note. Use # to separate the output diagnoses, then write a separate text that starts with *<*Reasoning*>* to explain the reasoning behind your answer: “. In addition to the prompt, context from the progress note was provided with the Subjective and Assessment sections. The Subjective section includes recent events during the hospitalization with relevant data, and the Assessment section includes the overall summary of the patient’s condition. Several examples were provided in a few-shot design from the training data to achieve full in-context learning for the LLM[20].

### Human Evaluation Framework for Diagnosis Generation

#### Development

The human evaluation was split into two parts: (1) Diagnostic Accuracy Evaluation; and (2) Diagnostic Reasoning Evaluation. The Diagnostic Accuracy part was benchmarked against the validated revised SaferDx instrument [21] that incorporates metrics for diagnostic error and safety. We defined a medical diagnosis using the Medical Subject Headings (MeSH) and National Cancer Institute (NCI) definitions from the National Library of Medicine Unified Medical Language System (UMLS). The MESH definition states, “The determination of the nature of a disease or condition, or the distinguishing of one disease or condition from another. Assessment may be made through physical examination, laboratory tests, or the like.” The NCI definition states, “The investigation, analysis and recognition of the presence and nature of disease, condition, or injury from expressed signs and symptoms; also, the scientific determination of any kind; the concise results of such an investigation.”

The diagnostic accuracy framework was categorized into the following components: (1) Overall Accuracy, (2) Plausibility, (3) Specificity, and (4) Omission/ Uncertainty. Accuracy, Plausibility, and Specificity were applied at the individual diagnosis level and were conditioned such that only diagnoses classified as accurate were scored for plausibility and only those classified as plausible were scored for specificity. Accuracy represented how well it met the definition of a diagnosis as stated above. Plausibility represented if the diagnosis was hallucinated and could pose potential harm. Finally, Specificity captured the level of detail in the diagnosis (i.e., sepsis vs sepsis from influenza pneumonia). The final component of diagnostic evaluation, Omission/ Uncertainty, was applied to the entire list of outputted diagnoses. Omission captured instances in which a diagnosis was missing from the output, but would be considered in a clinical setting. Uncertainty, which is conditional on Omission, further penalized a model for not utilizing the information it was provided versus not being provided with enough information.

The Diagnostic Reasoning Evaluation part was benchmarked against the framework established by Singhal et al. for evaluating the quality of evidence [22]. Their particular framework was supported by a high level of validity when compared to other established frameworks found during our literature review. The Reasoning Evaluation was categorized into the following components: (1) Comprehension, (2) Rationale, (3) Recall, and (4) Omission. In a similar manner to the Diagnostic Accuracy Evaluation, the Diagnostic Reasoning Evaluation was performed at the sentence level and the entire output level. Our goal in performing sentence-level evaluation as well as entire output was to increase inter-annotator agreement. Comprehension, Rationale, and Recall were pulled directly from the Singhal et al. framework and applied at the sentence level. Comprehension referred to reading comprehension and captured instances where the model did not understand information from the clinical note. Rationale captured instances in which the model took incorrect reasoning steps. Finally, Recall referred to the recall of knowledge and penalized a model for irrelevant, incorrect, or hallucinated facts. At the output level, Omission captured when a model failed to include explanations for diagnostic choices or conclusions.

The guideline with instructions for reviewers and implementation into the REDCap survey tool is shown in Supplementary Materials. In addition to the components outlined above, the guidelines present an additional component that was utilized in another project and not reported here. The questions for each component were scored using either a 5-point Likert scale or a binary decision. The Likert scales were Strongly Disagree to Strongly Agree except in the cases of Omission, Omission Uncertainty, and Specificity. The exact scales can be seen in Figure 3 and 4.

**Figure 3:**
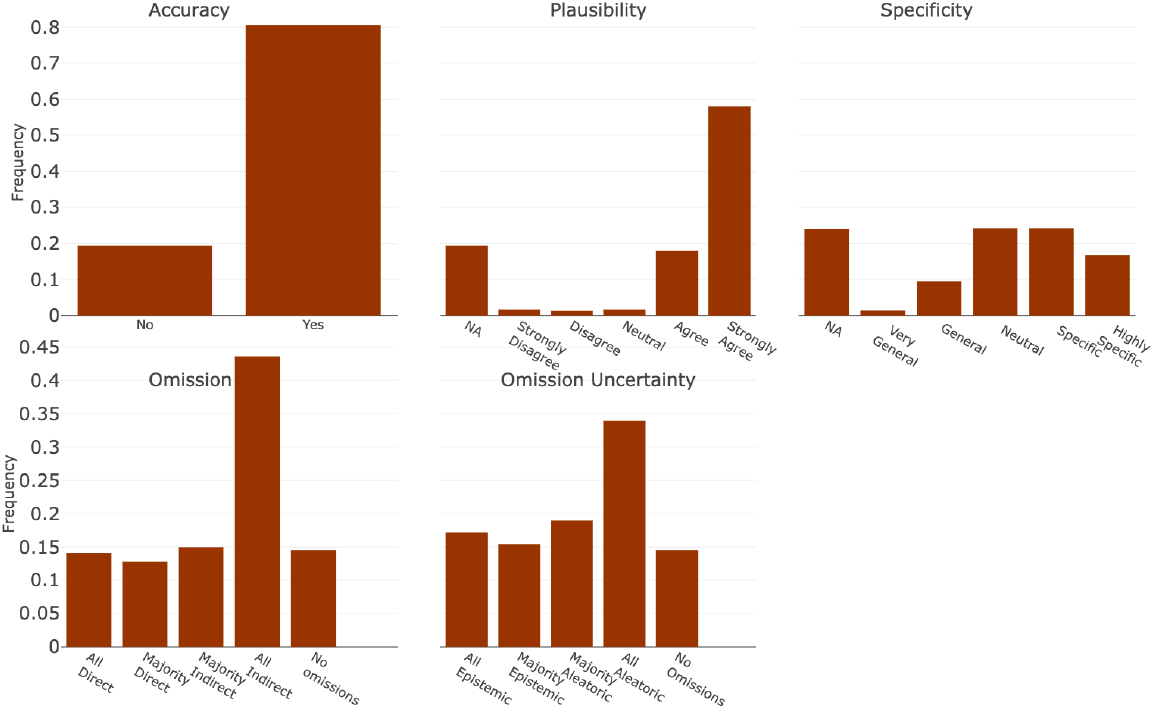
Diagnostic Accuracy Evaluation. Each bin refers to the text associated with the Likert Scale scoring scheme for each component in Diagnostic Accuracy Evaluation. They are represented in order where the far left bin represents a score of 1, or 0 in specific cases, and increases evenly by bin to 5.

**Figure 4:**
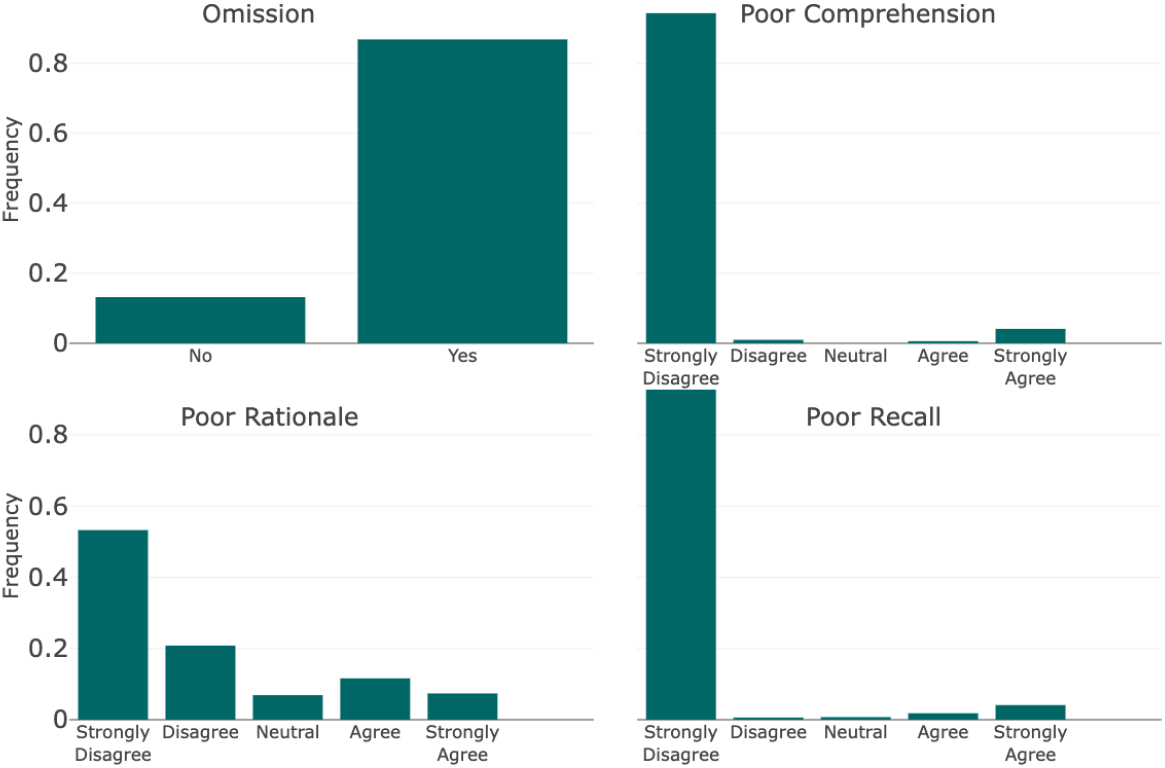
Diagnostic Reasoning Evaluation. Each bin refers to the text associated with the Likert Scale scoring scheme for each component. They are represented in order where the far left bin represents a score of 1, or 0 in specific cases, and increases evenly by bin to 5.

### Statistical Analysis of Human Evaluation Framework

The overall Diagnostic Accuracy and overall Diagnostic Reasoning scores were calculated using a linear combination of the components ascribed to each component. The scores were then normalized to a (0, 1) scale for a more direct comparison to the automated metrics. For every component, any NAs that were a result of branching logic were imputed to 0. The Comprehension, Rationale, and Recall scores were transformed (6 − *x*) to account for the inverse meaning of the questions on scoring. A Diagnostic Accuracy score (*D*_*i*_) for record ID *i* was therefore represented as

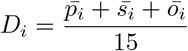

where for record ID i 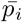 is the mean of the plausibility scores, 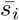 was the mean of the specificity scores, and *ō*_*i*_ was the mean of the omission and uncertainty scores. A Diagnostic Reasoning score (*R*_*i*_) for record ID *i* was similarly represented as

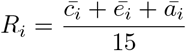

where for record ID i 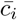 was the mean of the comprehension scores, ē_*i*_ was the mean of the recall scores, and ā_*i*_ was the mean of the rationale scores.

### Content Validity and Interrater reliability

The operating characteristics for the human evaluation framework were calculated by using standard definitions. Content validity was measured by first benchmarking the metrics to existing frameworks as described above and then reviewing the metrics in beta testing sessions with two senior physicians. Interrater reliability was assessed by comparing the evaluations of two senior physicians and two medical professionals against a predetermined threshold, necessitating adjudication until a kappa coefficient of at least 0.70 was achieved.

### Statistical Analysis of Automated Metrics against Human Evaluation

The ten automated metrics utilized in this analysis were organized into four categories: (1) N-gram overlap Based Metrics, (2) UMLS Based Metrics, (3) Non-UMLS Embedding Based Metrics, and (4) Learned Regression Based Metrics. There were three metrics in each category, except the n-gram overlap category which only had ROUGE-L. For COMET, we used the default model (wmt22-comet-da) and trained our model with the ProbSum training data. For the fine-tuned COMET model (ProbSum Trained COMET), we followed the training script outlined in the COMET github [23] using the default model as the base, the regression model configuration, and 20% of the ProbSum training data.

Only the Diagnostic Accuracy Evaluation was utilized from the human evaluation for correlation with the automated metrics. The Diagnostic Reasoning Evaluation was omitted since no ground truth labels were available. The Spearman, Pearson, and Kendall-Tau correlations were computed between each automated metric and the human evaluation scores. The Spearman and Kendall-Tau correlations both assume an independent, monotonic relationship between the compared populations, but the Kendall-Tau is more robust to small samples and many ties. The Spearman correlation uses the rank values of the cohorts, and the Pearson correlation uses the raw integer scores. Both assume linearity and normal distribution on two continuous populations. Additionally, we performed the Wilcoxon Signed Rank Test as a non-parametric method to assess the significance of the differences between the medians of two related samples between human metrics and automated scores, providing a deeper understanding. All correlation-related analyses and statistical testing were performed using R v4.3.1. Every automated evaluation metric was executed in Python 3.11.3.

## Results

### Human Evaluation

Given the input progress note without the Plan section that contained the ground truth diagnoses, ChatGPT generated 768 diagnoses with 828 reasoning sentences. Human evaluation was performed on every diagnosis and sentence. In human evaluation, ChatGPT had a median diagnostic accuracy score of 0.667 (IQR: 0.547 - 0.744), and a median reasoning accuracy score of 0.933 (IQR: 0.890-0.966).

In addition to the overall scores, the individual score components for the diagnosis and reasoning evaluations are presented in Figures 3 and 4, respectively. Among the diagnoses, 80.6% (n = 619), were classified as fulfilling the definition of a diagnosis, 75.9% (n=583) were deemed plausible, and 40.8% (n=313) were determined to be specific or highly specific. Of all the outputs generated, only 14.5% (n=27) had no diagnostic omissions and 14.0% (n=26) had omissions of diagnoses that were directly related to the input progress note. In addition, 32.5% (n=60) exhibited omissions from epistemic uncertainty.

In the reasoning score components, for every sentence scored, 94.3% (n=781) were deemed to have little to no evidence of incorrect reading comprehension. Furthermore, 92.7% (n=767) had little to no evidence of incorrect recall of knowledge and 74% (n=613) exhibited little to no evidence of incorrect reasoning steps. Across the reasoning paragraphs generated for the 186 progress notes, 13.1% (n=108) did not contain an explanation for the generated diagnoses.

### Automated Evaluation

The correlations between each selected automated metric and the corresponding human evaluation scores are shown in Table 1. The largest Spearman correlation with the human evaluation scores was with the SapBERT Score at 0.185. However, this correlation was not significantly different from many of the other correlations between an automated metric and human evaluation (p-value *>* 0.1). The largest Pearson correlation with the human evaluation scores was again with the SapBERT Score at 0.170. Similar to the Spearman correlation, this value was not significantly different from the other correlations, but the SapBERT Score was the only metric to have a 95% confidence interval that does not encompass 0 for the Pearson correlation coefficients. The final correlation metric, Kendall-Tau, showed the same trend as the other two. The Wilcoxon Signed Rank Test p-value was only favorable for the SapBERT Score at p=0.784 showing no difference between human evaluation and SapBERT. Overall, we did see a significant difference in correlations for the UMLS Based Metrics versus the Learned Regression Task-based metrics and some of the Non-UMLS Medical Embedding Based Metrics (p-values *<* 0.05). However, none of the automated metrics had a significant improvement over the ROUGE-L.

**Table 1:**
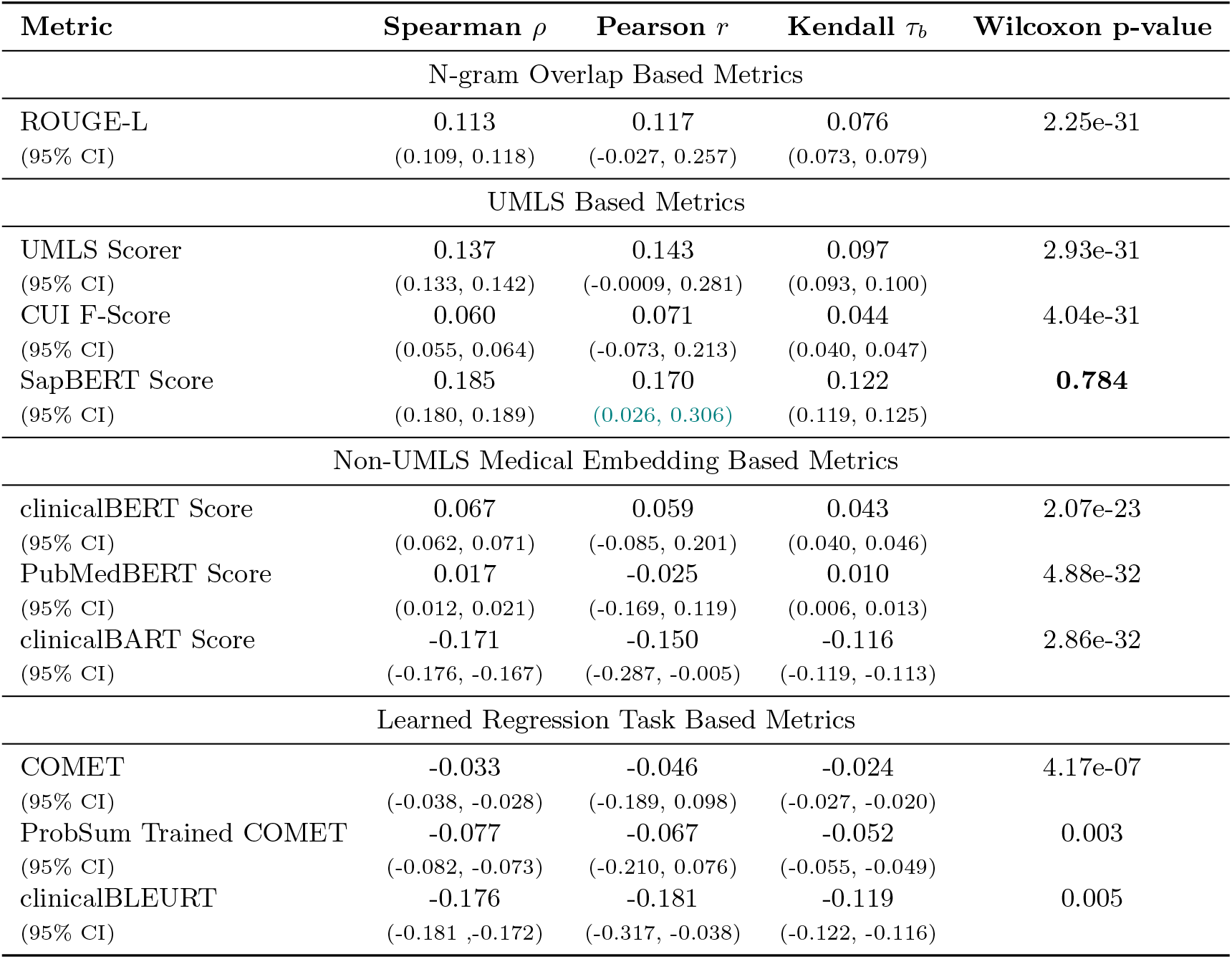
Correlation between Human Evaluation and Automated Metrics. Multiple correlation metrics and nonparametric paired statistical testing with Wilcoxon Signed Rank test for selected Automated Metrics and the Diagnostic Accuracy Evaluation from the Human Evaluation Framework. The 95% confidence intervals are listed in subscripts below each correlation.

| Metric | Spearman $\rho$ | Pearson $r$ | Kendall $\tau_b$ | Wilcoxon p-value |
| --- | --- | --- | --- | --- |
| N-gram Overlap Based Metrics |  |  |  |  |
| ROUGE-L<br>(95% CI) | 0.113<br>(0.109, 0.118) | 0.117<br>(-0.027, 0.257) | 0.076<br>(0.073, 0.079) | 2.25e-31 |
| UMLS Based Metrics |  |  |  |  |
| UMLS Scorer<br>(95% CI) | 0.137<br>(0.133, 0.142) | 0.143<br>(-0.0009, 0.281) | 0.097<br>(0.093, 0.100) | 2.93e-31 |
| CUI F-Score<br>(95% CI) | 0.060<br>(0.055, 0.064) | 0.071<br>(-0.073, 0.213) | 0.044<br>(0.040, 0.047) | 4.04e-31 |
| SapBERT Score<br>(95% CI) | 0.185<br>(0.180, 0.189) | 0.170<br>(0.026, 0.306) | 0.122<br>(0.119, 0.125) | **0.784** |
| Non-UMLS Medical Embedding Based Metrics |  |  |  |  |
| clinicalBERT Score<br>(95% CI) | 0.067<br>(0.062, 0.071) | 0.059<br>(-0.085, 0.201) | 0.043<br>(0.040, 0.046) | 2.07e-23 |
| PubMedBERT Score<br>(95% CI) | 0.017<br>(0.012, 0.021) | -0.025<br>(-0.169, 0.119) | 0.010<br>(0.006, 0.013) | 4.88e-32 |
| clinicalBART Score<br>(95% CI) | -0.171<br>(-0.176, -0.167) | -0.150<br>(-0.287, -0.005) | -0.116<br>(-0.119, -0.113) | 2.86e-32 |
| Learned Regression Task Based Metrics |  |  |  |  |
| COMET<br>(95% CI) | -0.033<br>(-0.038, -0.028) | -0.046<br>(-0.189, 0.098) | -0.024<br>(-0.027, -0.020) | 4.17e-07 |
| ProbSum Trained COMET<br>(95% CI) | -0.077<br>(-0.082, -0.073) | -0.067<br>(-0.210, 0.076) | -0.052<br>(-0.055, -0.049) | 0.003 |
| clinicalBLEURT<br>(95% CI) | -0.176<br>(-0.181, -0.172) | -0.181<br>(-0.317, -0.038) | -0.119<br>(-0.122, -0.116) | 0.005 |

In addition to correlations at the total score level, we also included correlations between each automated metric and a single component of the diagnostic score. Comparisons for the specificity component, plausibility component, and diagnostic omission component are outlined in Supplementary Materials. For specificity, the SapBERT Score had the highest Spearman correlation (*r* = 0.119) and the highest Kendall-Tau correlation (*τ*_*b*_ = 0.082) and the UMLS Scorer had the highest Pearson correlation (*ρ* = 0.135). For plausibility, the SapBERT score had the highest Spearman, Pearson, and Kendall-Tau correlations, 0.220, 0.180 and 0.154 respectively. Finally, for the diagnostic omission, the highest correlations for Spearman, Pearson, and Kendall-Tau were 0.109, 0.143, and 0.080 for the ROUGE-L automated metric. However, in each case, none of the correlations were significantly superior to the other automated metrics (p-value *>* 0.1).

## Discussion & Conclusions

This study introduces a human evaluation framework that offers a comprehensive assessment of essential elements critical to diagnostic evaluations in clinical decision support systems. Our findings reveal that numerous existing frameworks in the medical domain fail to include critical components such as content validity and inter-rater reliability, nor do they amalgamate various components into a single, simplified score for ease of comparison. Among the frameworks evaluated, the one proposed by Singhal et al. stands out as the most comprehensive, featuring a detailed revision system that ensures reliable evaluations. However, it primarily focuses on assessing the accuracy of medical facts within the MultiMedQA benchmark—a collection of six diverse medical question-answering datasets. This focus on question-answering tasks is common among current evaluation frameworks. In contrast, our framework extends beyond this limitation by offering a holistic evaluation of diagnosis generation, incorporating lessons from both the successes and shortcomings of previous methodologies. We have rigorously tested, reviewed, and validated our framework to guarantee a high-quality human evaluation standard for comparing automated metrics.

Our systematic review of the literature showcases the lack of automated evaluations that mirror human judgements in summarization tasks relating to the clinical domain. Each of the categories of automated evaluation was designed to leverage one component of natural language for evaluation. Despite expectations of strong performance from metrics categorized under Learned Regression Task-Based Metrics, these too exhibited a disappointing lack of correlation with human evaluations. This shortfall could likely be attributed to the relatively simplistic nature of the learning algorithms applied in our study. Furthermore, our iterations of these metrics, specifically tailored with clinical training using real-world patient progress notes, did not yield the anticipated improvement. In contrast, metrics based on the UMLS demonstrated some performance improvement over other metrics. Notably, SapBERT emerged as the most promising, benefiting from its training on the UMLS database. This allowed it to grasp subtle nuances by drawing on the extensive medical knowledge within UMLS. Its reliance on a cosine similarity score further minimized penalties for minor terminological discrepancies, provided the underlying semantics remained consistent.

These results highlight the central limitation of many automated evaluation metrics that rely on substandard references. The majority of automated metrics prioritize overlap and similarity, producing scores that may not accurately capture the factual accuracy or relevance of the text produced. Instead, the scores reflect the degree of structural and lexicographical resemblance between the generated text and the provided referents. While these elements remain important, they represent only a fraction of what constitutes a comprehensive evaluation. The quality of a reference text for evaluation can severely impact the reliability of an evaluative result. Moramarco et al. [24] highlighted this issue, pointing out the pronounced bias many automated metrics exhibit towards the structure of the reference text, as noted in their comparative analysis of numerous automated evaluation tools. This bias underscores the need for further research aimed at developing metrics capable of assessing the relevance of generated texts beyond mere semantic and lexicographical similarities. The challenge is particularly important in the medical domain, where generating diagnoses often involves navigating abstract concepts and addressing omissions—a scenario observed in over ten percent of our evaluations.

Similar work based on other shared tasks revealed similar conclusions. MedAlign [25], a dataset of instruction-answer pairs from electronic health records (EHR), found that COMET had the highest correlation with human judgments, but with a Kendall-Tau of 0.37 this correlation was only moderate. The MEDIQA 2021 shared task [26], focused on question and answer summarization of clinical text, did not compare any human and automated evaluation systems, but did show that across all three subtasks ROUGE-L and BERTScore showed a Pearson correlation of 0.409. Future work must be geared toward improving the state of evaluation to ensure that safety and accuracy remain at the forefront of improvement using generative AI in the clinical domain.

In conclusion, we introduced a human evaluation framework specifically designed to assess the key components of diagnosis generation relevant to diagnostic decision support systems, and we have shed light on the misalignment between human evaluations and automated metrics. Our framework prioritizes diagnostic safety and evidence-based reasoning, ensuring a thorough evaluation of diagnostic generation. We also focused on achieving a high degree of inter-annotator agreement. Our findings indicate that the automated metrics currently employed for this task fall short of expectations, as they fail to mirror human judgment accurately. However, we observed a notable improvement in UMLS-based approaches, attributed to their sophisticated representation of medical knowledge and their foundation in the extensive resources of the National Library of Medicine. This underscores the potential of leveraging medical knowledge databases to enhance the accuracy of automated evaluation metrics in the clinical domain.

## Data Availability

All data produced in the present study are available upon reasonable request to the authors

## Acknowledgements

We thank Anne Glorioso (UW-Madison) for their assistance with database selection and search query assistance as a computer science librarian. This work was supported by an NLM training grant to the Computation and Informatics in Biology and Medicine Training Program (NLM 5T15LM007359). YG was supported by K99LM014308 and DD, MA were supported by R01LM012973.

